# SARS-CoV-2 Testing Strategies for Outbreak Mitigation in Vaccinated Populations

**DOI:** 10.1101/2022.02.04.22270483

**Authors:** Chirag K. Kumar, Ruchita Balasubramanian, Stefano Ongarello, Sergio Carmona, Ramanan Laxminarayan

## Abstract

Although COVID-19 vaccines are globally available, waning immunity and emerging vaccine-evasive variants of concern have hindered the international response as COVID-19 cases continue to rise. Mitigating COVID-19 requires testing to identify and isolate infectious individuals. We developed a stochastic compartmentalized model to simulate SARS-CoV-2 spread in the United States and India using Reverse Transcriptase Polymerase Chain Reaction (RT-PCR) assays, rapid antigen tests, and vaccinations. We detail the optimal testing frequency and coverage in the US and India to mitigate an emerging outbreak even in a vaccinated population: overall, maximizing frequency is more important, but high coverage remains necessary when there is sustained transmission. We show that a resource-limited vaccination strategy still requires high-frequency testing and is 16.50% more effective in India than the United States. Tailoring testing strategies to transmission settings can help effectively reduce cases more than if a uniform approach is employed without regard to differences in location.

## 1. INTRODUCTION

Since the emergence of severe acute respiratory syndrome coronavirus 2 (SARS-CoV-2) in Wuhan, China in late 2019, the coronavirus disease (COVID-19) pandemic has resulted in more than 280 million reported cases and 5.4 million reported deaths worldwide as of January 1, 2022^1^. Despite efforts to curb the spread of SARS-CoV-2 through restrictions on travel^2^, business openings^3^, and personal measures^4^—including mask wearing and social distancing— cases have continued to rise in many countries^5^. Although vaccines against COVID-19 are available, the emergence of variants of concern^6^ that are only partially neutralized by existing antibodies or prior vaccination^7^ and are more contagious along with waning immunity^8^ has resulted in widespread COVID-19 outbreaks even in highly vaccinated populations^9^. Likewise, many populations, particularly in low- and middle-income countries, still lack widespread access to vaccines and continue to experience significant excess mortality^10^. All these factors must be simultaneously considered when developing mitigation strategies for emerging outbreaks. Consequently, it appears likely that SARS-CoV-2 will continue to pose a threat to public health for many years even if vaccines are distributed widely because of the rapid evolution of variants of concern. Thus, testing and containment will continue to be critical to COVID-19 response and mitigation.

Because of the high transmission rate of SARS-CoV-2 and prevalence of asymptomatic carriers^11^, accurate, efficient, and pervasive testing methods are needed to track and contain disease spread. Currently, two main diagnostic methods are widely used^12^. *Reverse Transcriptase Polymerase Chain Reaction* (RT-PCR), which is considered to be the gold standard, detects the presence of viral RNA in respiratory samples^13^. Although highly sensitive, test results typically require two to three days^14^, during which an infected individual may continue transmitting the virus. *Later flow rapid antigen tests* can be read in less than an hour^15^, and an individual who tests positive can immediately self-isolate. However, rapid antigen tests require a higher viral load for positive detection, must be tailored to the antigens on a specific variant, and may not return a positive result for infectious individuals past the early stages of their infection^16^.

Testing is only effective when individuals who test positive are quarantined, so they will not continue to transmit the disease further; essentially, testing and subsequent quarantining minimizes the effective duration during which an individual may spread the disease. However, testing usually only occurs in a reactionary fashion: for instance, an individual exhibits symptom that may be attributed to COVID-19 and the individual is tested, an individual comes in close contact with an infected person, or an individual seeks to travel and must show proof of a negative test. This type of testing is not optimized towards minimizing cases but rather just identifies current cases. Comparatively, proactive testing where testing is required within a community–such as with a college campus, business, or factory–is far more likely to be effective as it is more likely to capture asymptomatic spreaders.

Although both RT-PCR and antigen tests could play a role in COVID-19 containment, there is lack of clarity on the optimal coverage and frequency with which these tests should be used, and how they may be combined to greatest effect. Moreover, while COVID-19 has affected populations around the world regardless of socioeconomic status^17^, there are significant differences in transmission intensity by setting^18^. Tailored testing strategies that are specific to the local transmission context may be superior to applying a one-size-fits all method for all populations. Guidance on testing strategies may differ by transmission setting (i.e., in high-income countries, such as the United States, versus low- and middle-income countries, such as India) as not only do transmission dynamics differ^18^ but low- and middle-income countries (LMICs) now have the majority of confirmed COVID-19 cases^1^ along with more barriers to adequate health care^19^. Significant effort has been directed at containment strategies across high income countries in Europe and North America^20–24^, but these findings cannot be generalized to all countries worldwide^25^ because of differences in transmission intensity. Moreover, for COVID-19 to become globally endemic, it must be controlled in all countries, so determining setting-specific mitigation strategies and characterizing how vaccines and testing may be used in tandem is critical to international public health efforts to move towards the post-pandemic phase.

Here, we used a stochastic, compartmentalized, agent-based model (ABM) to simulate COVID-19 transmission dynamics in the United States and India under different proactive testing scenarios and vaccination potentials. We evaluated various strategies for COVID-19 control in both countries to identify their efficacy and costs and determine how testing recommendations may differ by transmission setting for mitigating an outbreak even in highly-vaccinated populations. Our simulation results can be directly applied to community settings, such as office buildings, factories, or campuses, that are trying to reopen safely and with minimal disease spread.

## 2. RESULTS

Before evaluating the effect of our testing scenarios, we ran our model without any mitigation (Fig. S1; Table S1) to validate our ability to recreate observed COVID-19 characteristics. The model-estimated case-fatality ratio (CFR) for the United States is 2.66% [95% percentile credible interval from 200 independent realizations of the ABM 2.17–3.12%], and the actual CFR at time of writing is 3.05%^26^; likewise, the model-estimated CFR for India is 1.85% [1.47–2.25%] and the actual CFR based on available data is 2.11%^18^. Since our model recreated the expected dynamics, we could justify the use of this model to identify the efficacy and trade-offs of testing strategies and vaccinations in both settings.

### Proactive Testing to Mitigate an Emerging Outbreak

The B.1.1.529 SARS-CoV-2 variant (designated “Omicron” by the World Health Organization) has shown almost complete evasion to existing vaccines or immunity from prior infection. Based on Omicron’s observed infectivity even in highly vaccinated populations, we modeled the impact of RT-PCR or antigen testing on a representative population in the US (Fig. 1) or India (Fig. 2) by proactively testing at various frequencies (quantified as days between tests) and coverages (quantified as the fraction of people surveilled each time testing is done).

**Figure 1:**
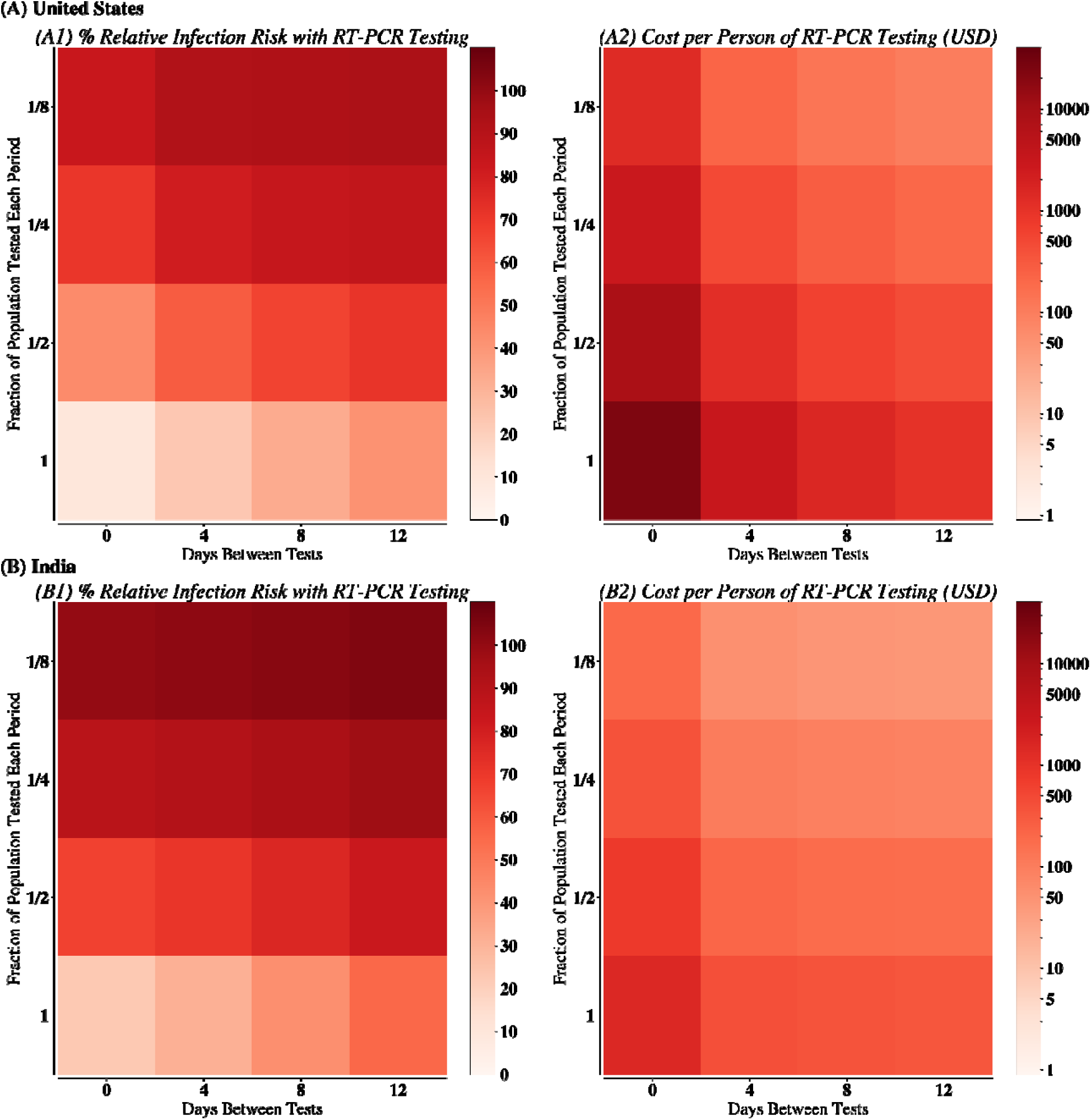
Relative infection risk and cost of testing for various RT-PCR scenarios in (A) United States and (B) India. (1) describes the relative infection risk of RT-PCR tests being used at the given frequency and coverage, compared with the scenario with no mitigation. (2) describes the cost of that testing scenario assuming that all susceptible individuals are tested at the appropriate frequency and coverage.

**Figure 2:**
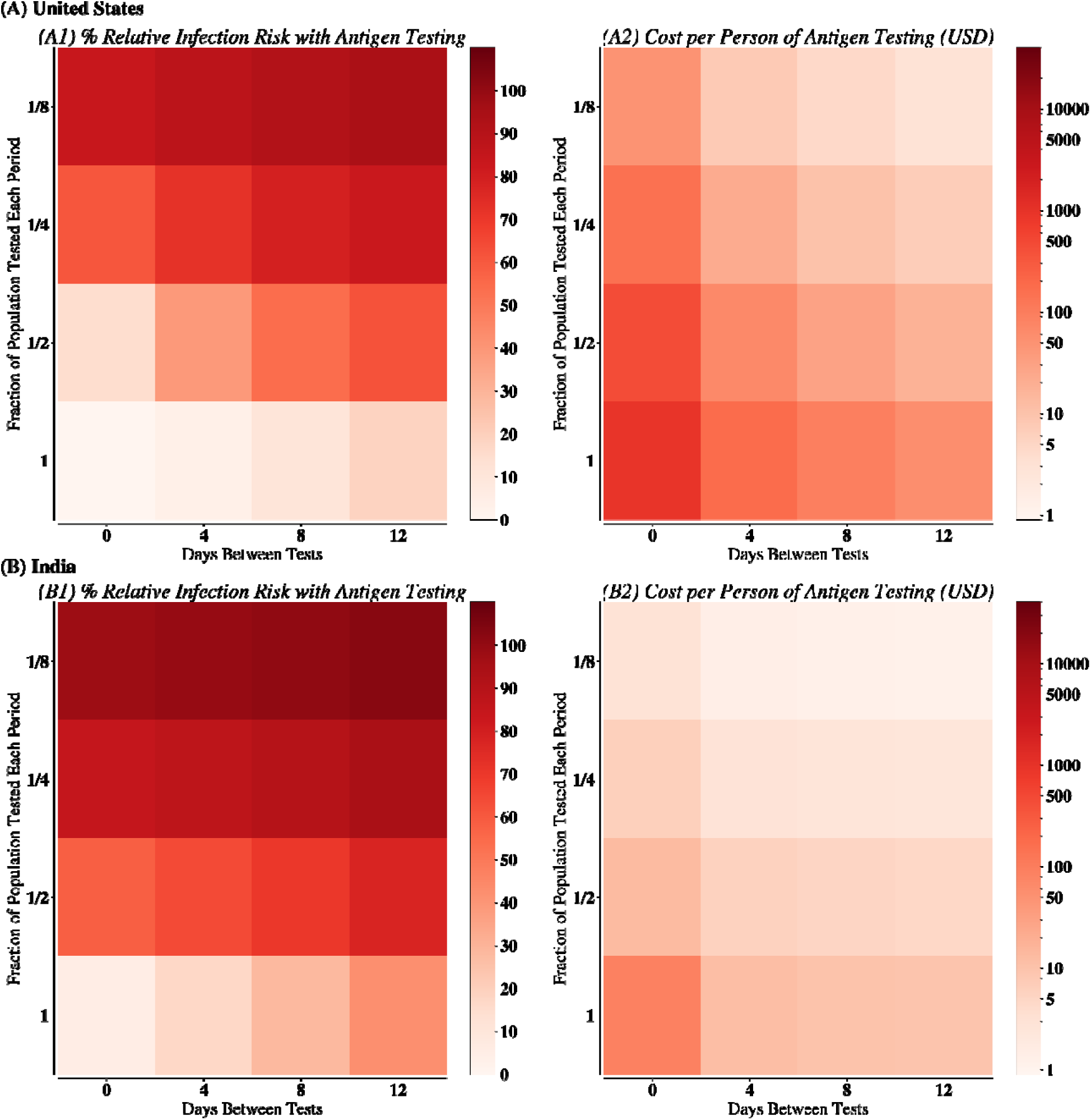
Relative infection risk and cost of testing for various antigen scenarios in (A) United States and (B) India. (1) describes the relative infection risk of antigen tests being used at the given frequency and coverage, compared with the scenario with no mitigation. (2) describes the cost of that testing scenario assuming that all susceptible individuals are tested at the appropriate frequency and coverage.

While the cost of the surveillance depended only on the rate of testing (i.e., average number of tests administered per day), the risk of infection depended on the frequency, coverage, type of test used, and transmission setting, with each test and setting placing different importance on frequency and coverage (Fig. S3).

Overall, for the same coverage and frequency of testing, using RT-PCR assays resulted in 12.65% [7.01% – 18.30%] more cases than antigen tests in the US and 9.30% [4.75% – 13.85%] more cases in India (Figs. 1-2), likely because RT-PCR tests require a two-to-three-day turnaround time during which infected individuals may continue transmitting (Fig. S2). Rapid antigen tests are also up to 20 times cheaper, allowing for further screening than RT-PCR assays with the same financial budget (Fig. 2). Ultimately, when used frequently and widely, antigen tests were notably better than RT-PCR assays (Fig. S3, S4, S5). Nevertheless, at low coverage and frequency, neither test was effective at mitigation.

Explanatory regression models that predicted the percent infected for each setting from frequency, coverage, and test used (*R*^*2*^ of all models >90%; Table S2) indicated that use of antigen tests versus RT-PCR tests resulted in 3.12% [95% confidence interval on the regression coefficients 1.73–4.51] fewer cases in the United States and 1.99% [95% CI 1.02–2.97] fewer cases in India. Independent of test, maximizing frequency had a larger effect on mitigating cases than maximizing coverage, with a 3.71-fold [2.99–4.43] greater effect in the United States and a 5.01-fold [4.15–5.87] greater effect in India. In other words, given resource constraints, the frequency of testing (i.e., the inverse of the number of days between subsequent test administrations) should be 3.71 and 5.01 times greater than the coverage (i.e., the number of testing administrations required to surveil the whole population) in the United States and India, respectively. Though frequency dominates overall, the importance of coverage differed by test. When RT-PCR tests were used rather than antigen tests, maximizing coverage has a 14.24% [10.71–17.76] greater effect on reducing cumulative infections in the United States and a 6.84% [4.89-8.79] greater effect in India. Nevertheless, use of antigen tests was more effective than use of RT-PCR tests, especially when used frequently.

While we observe the same general trends in both settings, frequency is more important in India than in the United States: Figs. 1 and 2 show a strong gradient of increasing cases as testing frequency is decreased, most noticeably for antigen testing and especially for India. However, we observe that at low coverage, the effect of frequency is much less in India. High-frequency, low-coverage testing can still be useful in the United States, but we did not observe the same pattern in India, where coverage must be relatively high for effective mitigation. Although increasing coverage when it was low had little benefit, increasing coverage from half of the population surveilled to the whole population surveilled had a larger effect in India. Ultimately, though frequency may still have dominated overall, increasing coverage was also critical in certain testing scenarios in India. Additionally, we conducted sensitivity analyses by running canonical strategies with R_eff_ from 2.5 (used in all main figures and analyses; Fig. S6, Fig. S7) and decreasing R_eff_ to R_eff_ = 2 (Fig. S8) and 1.5 (Fig. S9). Our main findings and trends still held across all values of R_eff_, though there are fewer cases with a lower R_eff_.

### Utilizing Combinations of Tests and Vaccines

Emerging variants of concern are not only vaccine-evasive but often more transmissive. To identify and isolate transmissive individuals as quickly as possible, we propose a mixed strategy that utilizes the complementary nature of antigen tests and RT-PCR assays (i.e., antigen tests excel when used frequently whereas RT-PCR assays are more effective when used widely). All individuals were tested weekly using antigen tests and all negative results were followed up immediately by an RT-PCR assay (Fig. 3). Since RT-PCR assays have higher sensitivity, their use as a follow-up should allow for the detection of individuals with a viral load too low to be detected by an antigen test. Nevertheless, this was resource intensive: it potentially required more than just one test per individual.

**Figure 3:**
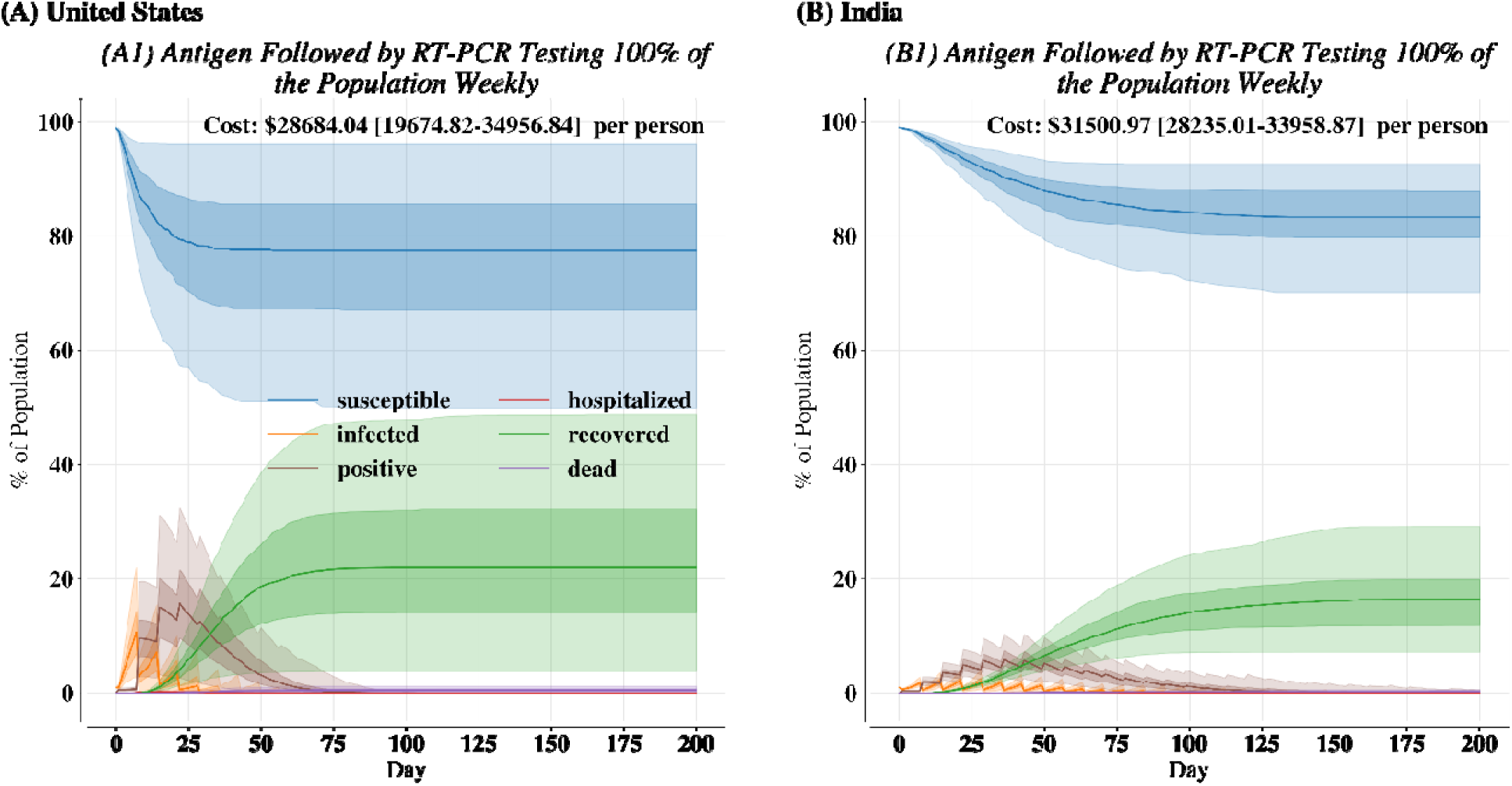
Disease course in (A) United States and (B) India for antigen testing followed by RT-PCR testing of antigen-negative individuals 100% of the population weekly. Bold lines are the median over 200 independent replicates. Dark-shaded regions are th 25^th^ to 75^th^ percentiles. Light-shaded regions are 2.5^th^ to 97.5^th^ percentiles.

Such a mixed strategy was effective in both settings—resulting in minimal hospitalizations and deaths—but more effective in reducing cases India than in the United States. Compared with weekly antigen testing (Fig. S7), this approach did not significantly reduce cases in the United States (*λ =* 2.25, *p =* 0.13) or India (*λ* = 1.00, *p* = 0.32). Nevertheless, this strategy resulted in reaching 1% incidence 7.0 [0.0–14.0] (*λ =*9.25, *p* < 0.005) days earlier in the United States and 7.0 [–7.0–7.0] days earlier (*λ = 5*.*81, p* = 0.016) in India. In practice, effective mitigation of SARS-CoV-2 or conferring immunity through vaccination or booster shots can allow testing to slow down and be less extensive, decreasing the required cost of testing. However, testing should only stop after SARS-CoV-2 infections have dropped below a level where they are no longer able to trigger widespread transmission or enough individuals are vaccinated (such external factors are not considered in our simulations as they are highly variable and differ between transmission setting). Nevertheless, we show that effective surveillance is critical especially in the early stages of transmission to mitigate disease burden from contagious variants of concern.

Likewise, the rapid emergence of variants of concern has placed an emphasis on widespread vaccination campaigns and now even booster shots for some populations. Nevertheless, key questions remain about optimal testing strategies and epidemic trajectories for populations as vaccines are administered or additional immunity is conferred through booster shots. Thus, we determined how proactive testing and simple vaccination strategies can be used together to promote SARS-CoV-2 reaching the endemic phase. Disease surveillance, continued testing, and vigilance remain critical even as more individuals become vaccinated. Note that while there are studies detailing optimal vaccine allocation strategies^27^, our goal is not optimize the distribution of vaccines but rather determine the impact of vaccines in a resource-limited distribution scheme and how vaccines must be coupled with testing to mitigate disease spread. We couple daily vaccinations with antigen testing 100% of the population weekly and antigen testing 33.3% of the population weekly (i.e., see Fig. S7 for these scenarios but without vaccines) in both settings.

Overall, vaccines are mostly effective in reducing COVID-19 cases at both coverages and in both transmission settings. First, compared to there being no vaccinations or immunity, 20% vaccines coupled with weekly antigen testing 100% of the population resulted in 15.12% [6.39 – 25.50] (*λ = 396*.*01, p* < 10^−6^) fewer cases in India than without vaccines. However, surprisingly, the limited distributions of vaccines only reduced cases marginally in the US compared to weekly antigen testing alone (*λ = 1*.*69, p* = 0.19). Secondly, we observe that the increasing number of vaccinations resulted in a mitigated disease course; concretely, we observed that vaccines coupled with weekly antigen testing 100% of the population resulted in infections below 0.5% incidence 7.0 [0 - 15.0] (*λ = 45*.*00, p* < 10^−6^) days faster in the US and 70.0 [34.83 – 126.0] (*λ = 372*.*49, p* < 10^−6^) days faster in India than the same testing strategy but with no vaccines. However, widespread testing is still critical, especially in early phases of vaccine distribution where we note many antigen-positive SARS-CoV-2 cases being captured. We also found marked differences between testing 100% of population weekly or 33.3% of the population weekly: specifically, testing 100% of the population weekly resulted in 30.83% [20.41 – 33.10] (*λ = 342*.*25, p* < 10^−6^) fewer cases in the US and 12.99% [7.65 –18.58] (*λ = 396*.*01, p* < 10^−6^) fewer cases in India than testing 33.3% of the population weekly, suggesting high frequency testing remains critical even with vaccines.

Nevertheless, while vaccines are effective in both transmission settings, there are key differences in the corresponding disease courses and number of infections by transmission setting. In particular, as discussed above, cases peak earlier in the US than in India, and as a result of distributing vaccines while there is still transmission in India, there were 16.50% [3.83 – 33.17] (*λ = 396*.*01, p* < 10^−6^) fewer cases in India than the US for antigen testing 100% of the population weekly. Furthermore, we also noted that using vaccines reduced the cost of testing because vaccinated individuals are not tested. Ultimately, the use of vaccines is effective in minimizing cases when coupled with testing and can be a tailored strategy to mitigate ongoing outbreaks.

## 4. DISCUSSION

### Maximizing the Effectiveness of Testing Strategies

Our results provide insight into constructing testing strategies with maximum effectiveness. First, antigen tests are more effective than RT-PCR tests across both transmission settings because they enable faster action to reduce transmission; our results agree with real-world evidence that antigen tests have been used successfully in nation-wide testing campaigns^28,29^. We observe that RT-PCR assays are more comparable to antigen tests as their turnaround time is decreased^30^ (Fig. S2). Nevertheless, the increased mitigation of antigen tests compared with RT-PCR assays with standard turnaround times is most pronounced when 100% of the population is tested weekly. Since antigen tests have a quicker turnaround time, infected individuals are more likely to self-isolate faster. Our simulations show that use of antigen tests results in a lower peak of daily cases compared with RT-PCR assays. Because disease spread is greatest in the early stages, when most of the population is still susceptible, early isolation of infected individuals is critical to mitigating disease spread^31^, especially critical when considering highly contagious and vaccine-evasive variants of concern. Additionally, SARS-CoV-2 transmission to secondary individuals is significant even immediately after initial infection^32^ (Fig. S9), further underscoring the need for isolating infectious individuals quickly.

We show that that high-frequency testing must be prioritized when fighting an emerging outbreak driven by a contagious variant of concern, though the relative importance of frequency versus coverage differs by setting and the type of test used. Maximizing frequency has the greatest importance for antigen testing. This is likely driven by its lower sensitivity but quicker turnaround: since antigen tests are unable to detect infected individuals with low viral loads, they must be used frequently to identify when individuals become infectious past detectable levels and force them to isolation. On the other hand, RT-PCR assays can still be effective when used widely because they can identify infectious individuals with low viral loads. Moreover, frequent use of antigen tests is not substantially better than even more extensive disease mitigation strategies, such as coupling antigen and RT-PCR tests, and is still effective in scenarios where there may even be unmitigated disease spreaders.

### Comparison of Testing Effectiveness between the United States and India

Although we observe mostly similar trends in mitigation strategies between the two countries, some differences are important for tailoring mitigation solutions. First, while both transmission settings have the same R_eff_ and are parametrized with the same incubation period and transmission distribution, the disease trajectory is markedly different. Overall, transmission is shorter, reaches a higher peak in percentage infected, peaks earlier, and the credible intervals are substantially larger in the United States than India—all of which can be explained by the contact matrices and the inherent variability in contact patterns^18^; moreover, our findings generally agree with real world analyses of SARS-CoV-2 spread in the US^33,34^. Notably, the large credible intervals in the United States are likely driven by the high variability in the distribution of infected secondary contacts^35^. Moreover, these differences affect mitigation effectiveness. We note that sustained transmission in India, which our simulation predicts, is similar to what is actually occurring in India and contributing to observed resurgences of cases^36^.

While our simulations overall indicate that high-frequency testing must be an urgent priority, we also find that the importance of frequency and coverage differs by transmission setting. Whereas increasing frequency is overall more important in India, increasing coverage beyond half of the population surveilled at each testing occurrence is critical for markedly improved mitigation and for the benefits of frequency to be most noticeable. Notably, at lower coverages, increasing frequency is more beneficial than increasing coverage. Consequently, we suggest that with limited resources, frequency should be prioritized unless coverage can be increased beyond half of the population surveilled; likewise, at those high coverages, the importance of frequency is most evident. Ultimately, the need for widespread and frequent antigen testing is urgent in both countries, but the trade-off between frequency and coverage should be tailored to community needs.

Moreover, we find antigen testing is not only more effective but also substantially cheaper than use of RT-PCR assays. Our simulations show that given a constant budget constraint, antigen testing can be done more frequently or at wider coverage and result in fewer cases than use of RT-PCR assays. Nevertheless, we also observe that the same testing scenario may have different costs in the United States versus India. In our simulation, we assume that all individuals who have not been infected must be tested. Since in the United States the peak in cases occurs earlier, more individuals are infected in the early stages and thus a typical individual is removed from the testing pool faster than in India. Although the cost of testing thus should be lower in the United States and we do observe this in many of our simulations, in certain scenarios (e.g., where antigen tests are used at high frequency and coverage; Fig. 2), the cost is less in India because the testing strategy is less effective. Since in India more individuals are infected and do not need to be tested, the cost of the strategy falls. However, given the differential nature of disease spread, testing frequency and coverage can change as the epidemic progresses, which may also change the cost (not considered in our simulations). Finally, we do not consider the cost of hospital beds or self-isolation, which likely differ heavily between settings. Additionally, our analysis does not explicitly consider contact tracing or self-isolation of individuals who experience symptoms, so our results more directly indicate the impact of proactive testing and immediate quarantining.

Finally, we show that vaccines and testing can be combined to create mitigation strategies that mitigate the duration of sustained transmission and can usher in an endemic phase earlier. Even a resource-limited vaccine allocation strategy of simply distributing vaccines randomly to susceptible individuals in addition to testing some of the population weekly is effective in minimizing cases and ending sustained transmission earlier in both transmission settings (Fig. 4). However, we show that vaccinations have different impacts in each transmission setting. In particular, the vaccination strategy utilized is more effective in reducing cases in India than the US. This is likely due to the sustained nature of SARS-CoV-2 transmission observed naturally in India (Fig. S1); consequently, the impact of continued vaccinations is greater in India as supposed to the US where infections peak much earlier. Thus, our findings show that vaccines are critical to minimizing the chance of future waves of COVID-19 cases especially as much of the world’s population still remains susceptible to SARS-CoV-2. Nevertheless, we note that widespread testing is still critical, especially in the early phases of vaccine distribution when vaccines are limited. Moreover, testing will likely continue to be critical as further variants of concern that may be vaccine-resilient or even vaccine-resistant continue to emerge and must be monitored to ensure that resurgences of SARS-CoV-2 infections do not occur^37^.

**Figure 4:**
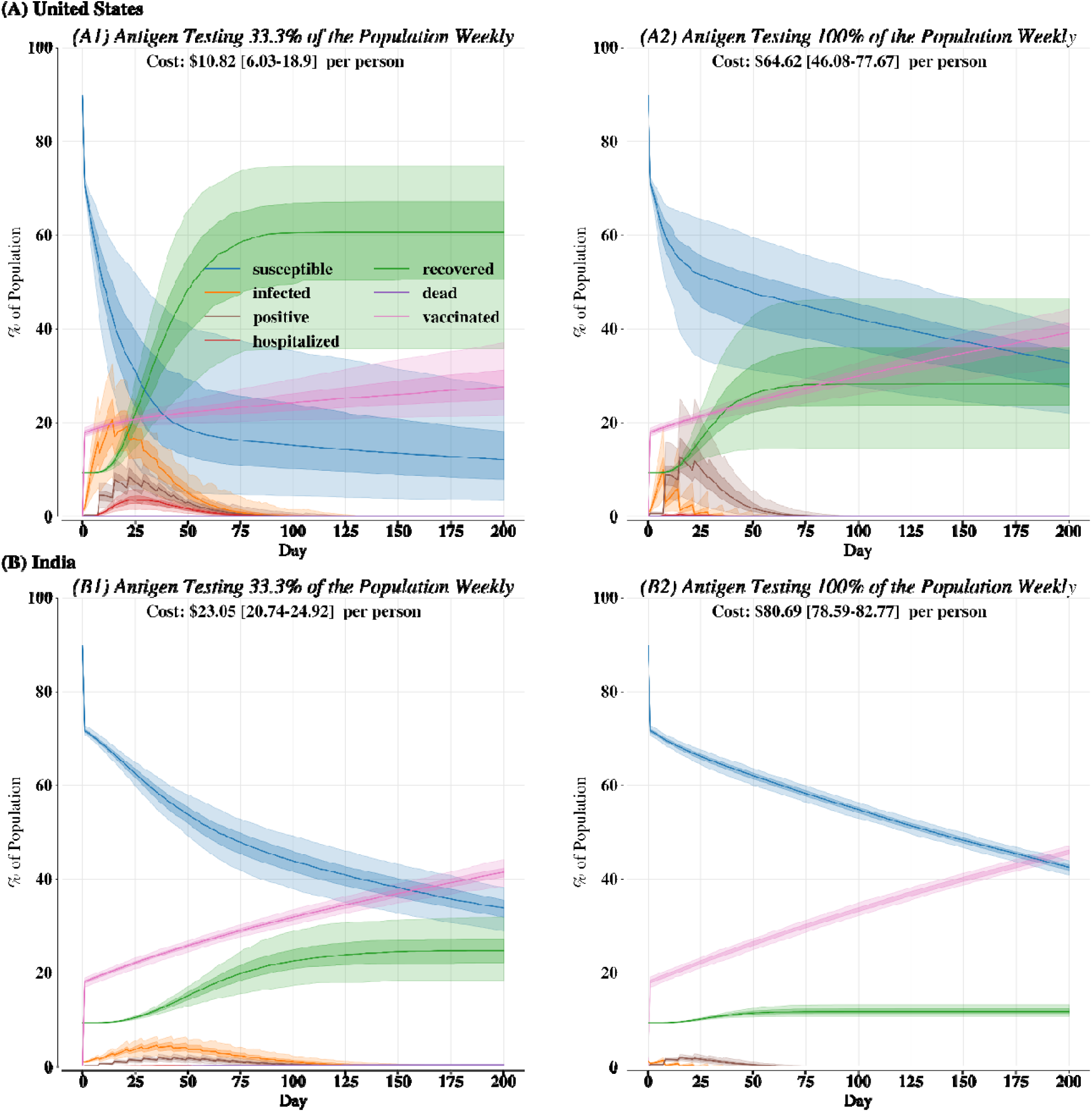
Disease course for (A) United States and (B) India with 20% of the population as initially vaccinated and then 0.25% of the susceptible population being vaccinated each subsequent day; vaccines are coupled with antigen testing (1) 33.3% or (2) 100% of the population weekly. Bold lines are the median over 200 independent replicates. Dark-shaded regions are the 25^th^ to 75^th^ percentiles. Light-shaded regions are 2.5^th^ to 97.5^th^ percentiles.

Generally, the most effective of the testing strategies discussed in this paper are frequently not the most expensive but rather those that are most closely tailored to the dynamics of the setting. Therefore, identifying transmission dynamics across a wide range of settings and applying specialized testing scenarios to specific environments are critical to effective mitigation. Our study suggests that contact matrices specific to the setting must be used as opposed to generic contact matrices^38^ commonly used in modeling studies. Our simulation shows that social mixing patterns affect the efficacy of mitigation strategies. However, we acknowledge that in developing tailored scenarios, considering social factors is also crucial, since health behaviors have been shown to be related to social clustering^39–41^. Nevertheless, we believe the trade-offs presented in this paper present a useful set of heuristics that can inform testing strategies and health policy across a wide range of settings.

## 5. METHODS

Accurately simulating SARS-CoV-2 dynamics requires two types of data: details of disease outcomes (i.e., data regarding COVID-19’s effects in humans), and attributes of virus spread (i.e., properties of SARS-CoV-2 transmission). Details of disease outcomes include such effects as whether an infection will result in hospitalization. Attributes of virus spread refer to the disease’s epidemiology, such as the Reff and viral shedding by day. We gathered both kinds of data through freely and publicly available sources for both transmission settings.

### COVID-19 Characteristics

We obtained data on COVID-19 cases and death counts from the beginning of data acquisition in the United States and India: US data were obtained from the National Center for Health Statistics (of the US Centers for Disease Control and Prevention, CDC)^26^, and data from India were obtained from previous studies^18,42^. We assign each individual an age, gender, and comorbidity based on census data and comorbidity prevalence^43^. We used these data to estimate COVID-19 probability of death given age, gender, and comorbidities. Additionally, the CDC keeps data on the probability of hospitalization and summary statistics (i.e., the 25^th^, 50^th^, and 75^th^ percentile) for time spent in the hospital due to severe COVID-19 infection^44^. Although having the distribution of hospital stay duration would be ideal, based on empirical evidence that such data follow a negative binomial distribution^45^, we construct negative binomial distributions with the same summary statistics to generate an estimated probability for various lengths of hospital stays. We had different hospitalization rates in the United States and India, but we assume that the pattern of hospital stay length is maintained between the two countries, since there is a shortage of data on COVID-19 hospitalizations in India. Ultimately, drawing from empirical evidence of COVID-19 effects, we project the expected outcome of individuals’ infection given their age, gender, and comorbidities, including whether they are hospitalized, time spent in hospital, and whether they die.

### SARS-CoV-2 Tests

We considered two types of tests in our simulations: RT-PCR assays and antigen tests. The sensitivity and specificity as a function of viral load of each test are given in Table S3. From current data, we used a turnaround time of three days for RT-PCR assays^14^. We assumed antigen test results come back quickly enough that an infectious individual will not further spread the virus while waiting for results. Finally, drawing from current estimates of costs for these tests, we assumed a cost of each test (Table S3), but note that costs may change dramatically based on setting and health insurance coverage. All individuals who do not yet have a documented infection are tested. Likewise, based on current evidence, fully vaccinated individuals (i.e., either those with a two-dose regiment of an mRNA vaccine or a single dose of another WHO approved vaccine) experience a decreased probability of transmitting the disease, and further booster shots increase the chance that a vaccinated but infected individual will not transmit to their contacts.

### SARS-CoV-2 Epidemiology

Although the above data determine the effects of COVID-19 for an individual, they do not detail how SARS-CoV-2 spreads in a population. Thus, to simulate SARS-CoV-2 transmission, we gathered data on the incubation period^46^, transmission probability since infection^32^, and inferred viral load after symptoms^47^. Together, these variables detailed the necessary information for SARS-CoV-2 transmission from an infected individual to secondary contacts by day and whether the individual will be flagged as infectious by a test. Despite extensive data along with quantifiable uncertainty for the incubation period, transmission probability since infection, and viral load after symptom onset of SARS-CoV-2 (see Table S3 for the parameters used in our analysis), quantifying viral load prior to symptom onset is more difficult and there is sometimes contradictory evidence on the peak viral load^48,49^. Thus, we drew from previous viral kinetics models to infer the viral load prior to symptom onset; note that our results are likely robust to changes in viral load distribution as there is variability in our estimated viral loads by individual (Fig. S9). Specifically, we say that viral load peaks anywhere from day 0 to day 4 after symptom onset^50^; the peak is anywhere from 5 to 11 log10 virions per mL^47^, that log10 viral load increases linearly from negative infinity on the day of infection to the aforementioned peak, and that log10 viral load decays from peak to the end of the individual’s infection linearly with a slope drawn from meta-analyses^47^. See Fig. S9 for our inferred viral load distributions.

Additionally, we gathered data on contact matrices and the distribution for the number of secondary cases arising from an infected individual for each transmission setting^18,51^. These variables were used for determining how many infections may arise from a single infected individual and the likely age of the consequently infected individuals. Table S3 shows a complete list of parameters compiled to simulate SARS-CoV-2 transmission in our model. From these parameters, we simulated realistic disease spread in a population across settings.

### Model Description

We developed a stochastic, compartmentalized, empirically driven agent-based model (ABM) to project COVID-19 cases, hospitalizations, and deaths given a variety of testing strategies. We adopted the following structure in our model: individuals in the population start as “susceptible” or “recovered” if they have previously been infected before entry into our simulation. Susceptible individuals can become “infected and not expressing symptoms” after a positive transmission event with another infected individual. Individuals can either stay as “infected and not expressing symptoms” (i.e., “asymptomatic”) for the duration of their infection or move to “infected and expressing symptoms”. Infected and symptomatic individuals may either recover or become hospitalized. Finally, hospitalized individuals may either recover or die (see Table S3 for the probability and duration of each event). Throughout each phase, the individual’s probability of transmitting the virus changes (peaking near symptom onset), as does the viral load (peaking shortly after symptom onset). We inferred the viral load before symptom onset based on previous studies^30^ and drew the viral load after symptom onset from meta-analyses^47^. Nevertheless, not all individuals will transmit the virus: in accordance with “superspreading”^52^, we drew the number of positive contacts for infected individuals from a negative binomial distribution, and whom they are likely to infect, from contact matrices. Individuals interacted homogeneously with each other in Brownian fashion in an open space with dimensions tuned to ensure the R_eff_ is 2.5 without any mitigation. ABMs present two benefits over traditional deterministic compartmentalized models: (i) implementing individual specificity is easier, and (ii) they are inherently stochastic and thus can provide credible ranges of the epidemic trajectory given initial conditions. Each model was run in the following way: there are 5,000 individuals with age and genders drawn from US and Indian census data, and comorbidities drawn from recorded prevalences given age and gender in 2017. Note that these parameters can be easily changed so that policymakers can determine which mitigation and testing strategies are most effective for specific communities. We ran each model for 200 days (until a steady state is reached), 200 times (i.e., independent replications), and present the 50th, 2.5th, 25th, 75th, and 97.5th percentiles (i.e., the “credible intervals”) as summary statistics in figures; when reporting values in the main text, we simply provide the 50th percentile and the 2.5th and 97.5th percentiles as the credible interval. We use standard methods of propagating uncertainty in variables to ensure meaningful credible intervals^53^. Unless otherwise mentioned, to test for statistically significant difference in medians between two distributions, we use a two-sided paired Mood’s Median test^54^. The model was developed in Python^55^ using the Mesa package under the Apache2 license^56^. All subsequent analyses were done in Python.

## Data Availability

All data used in this paper is freely and publicly accessible through the US Center for Disease Control or peer-reviewed studies. The provided supplementary materials document details data sources and includes citations to all the relevant sources.

## ACKNOWLEDGEMENTS

The authors are grateful for the computational resources managed and supported by Princeton Research Computing, which were used for all simulations in this paper. This work was supported by the Foundation for Innovative New Diagnostics.

## AUTHOR CONTRIBUTIONS

R.L. conceived the study. C.K. and R.B. developed the models and performed the analyses. R.L., S.O., and S.C. provided validation of study findings. C.K. and R.B. prepared the first draft of the manuscript; all authors contributed to the revised manuscript.

## DATA AVAILABILITY

All data used in this paper is freely and publicly accessible through the US Center for Disease Control or peer-reviewed studies. The provided supplement details data sources.

## CODE AVAILABILITY

The code used in this paper is available upon request and is on <u>GitHub</u>.

