## Supplementary Materials for "SARS-CoV-2 Testing Strategies for Outbreak Mitigation in Vaccinated Populations"

Supplemental Materials

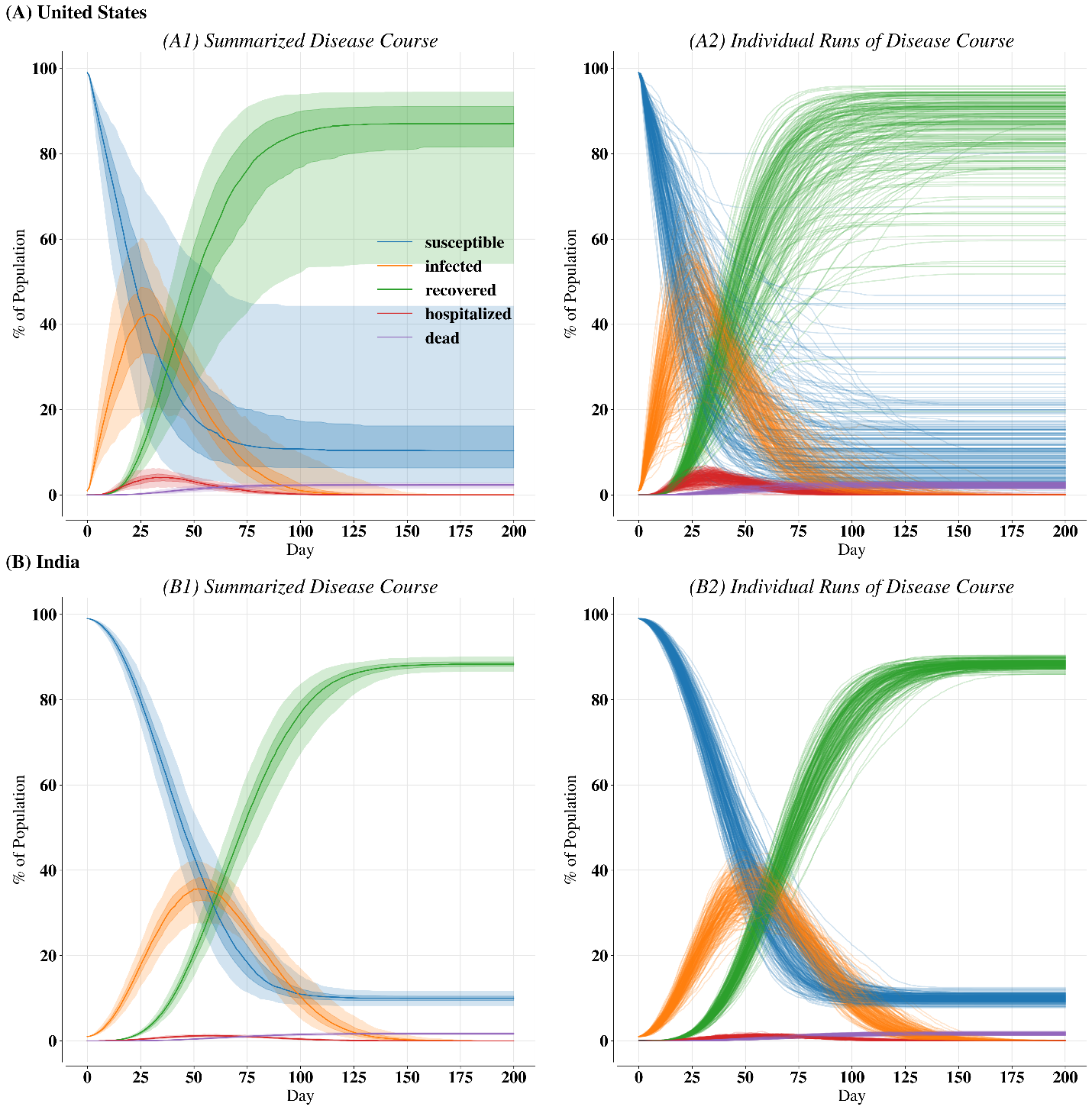

Figure S1: Disease course with no mitigation for (A) the US and (B) India. Left plots are the summarized disease course where bold lines represent the median of 200 individual runs of the IBM, dark shaded areas represent the 25^th^ to 75^th^ percentile of those 200 runs, and light shaded areas are the 2.5^th^ to 97.5^th^ percentiles. Right plots are the individual runs.

Table S1: Percent of individuals infected [95^th^ percentile credible interval from 200 independent runs of the model] for the exploratory testing scenarios in Fig. 1.

|  | RT-PCR | | Antigen | |
| --- | --- | --- | --- | --- |
| % of population tested weekly | 33.3 | 100 | 33.3 | 100 |
| % infected in US | 84.22 [34.164 – 93.234] | 71.19 [29.018 – 87.774] | 67.65 [19.389 – 88.388] | 20.09 [4.820 – 48.936] |
| % infected in India | 85.52 [82.916 – 87.681] | 74.28 [65.596 – 78.883] | 80.75 [75.623 – 83.562] | 17.50 [7.733 – 29.502] |

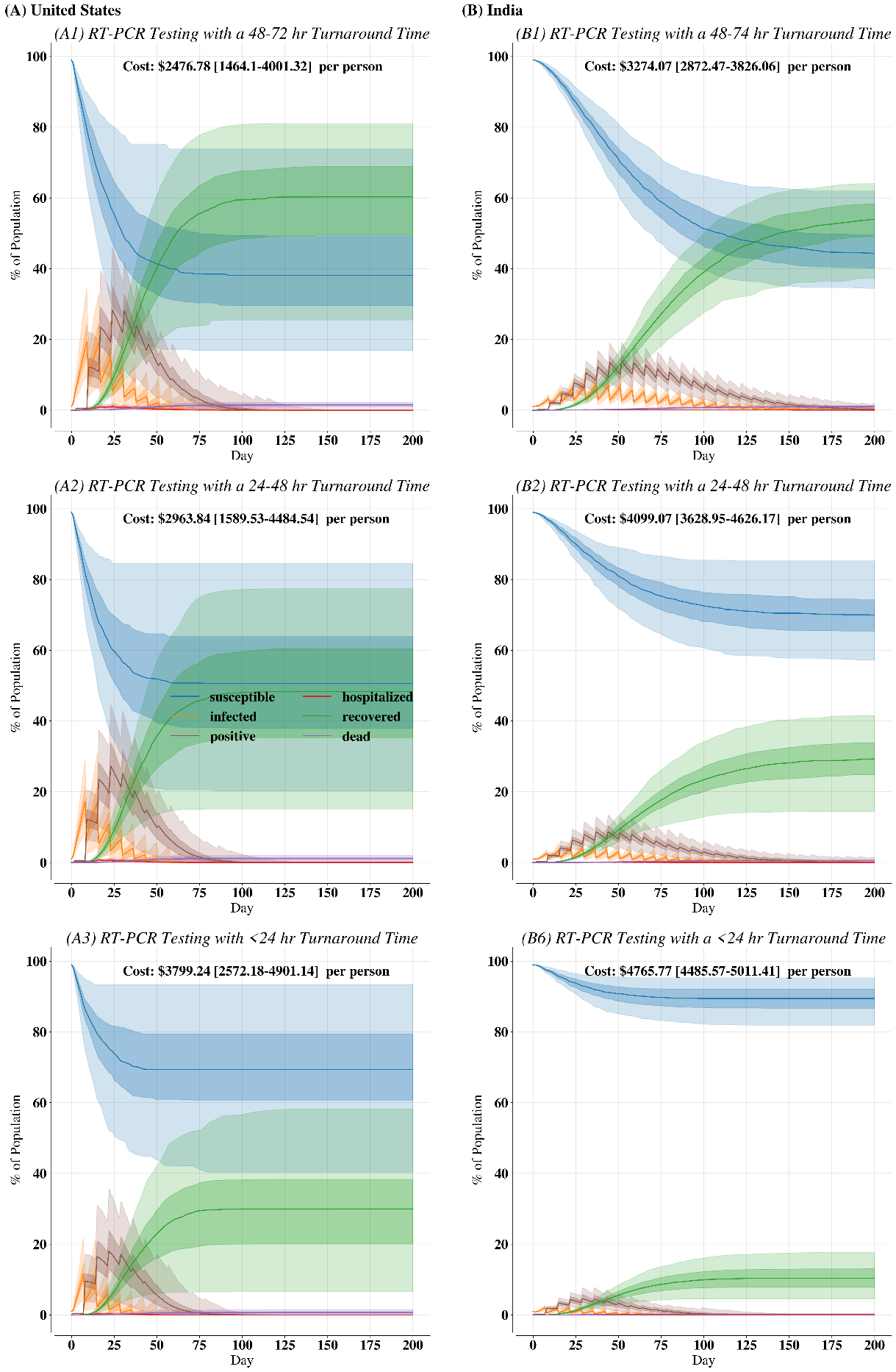

Figure S2: The impact of turnaround times in RT-PCR testing for (A) the US and (B) India when 100% of the population is RT-PCR tested weekly.

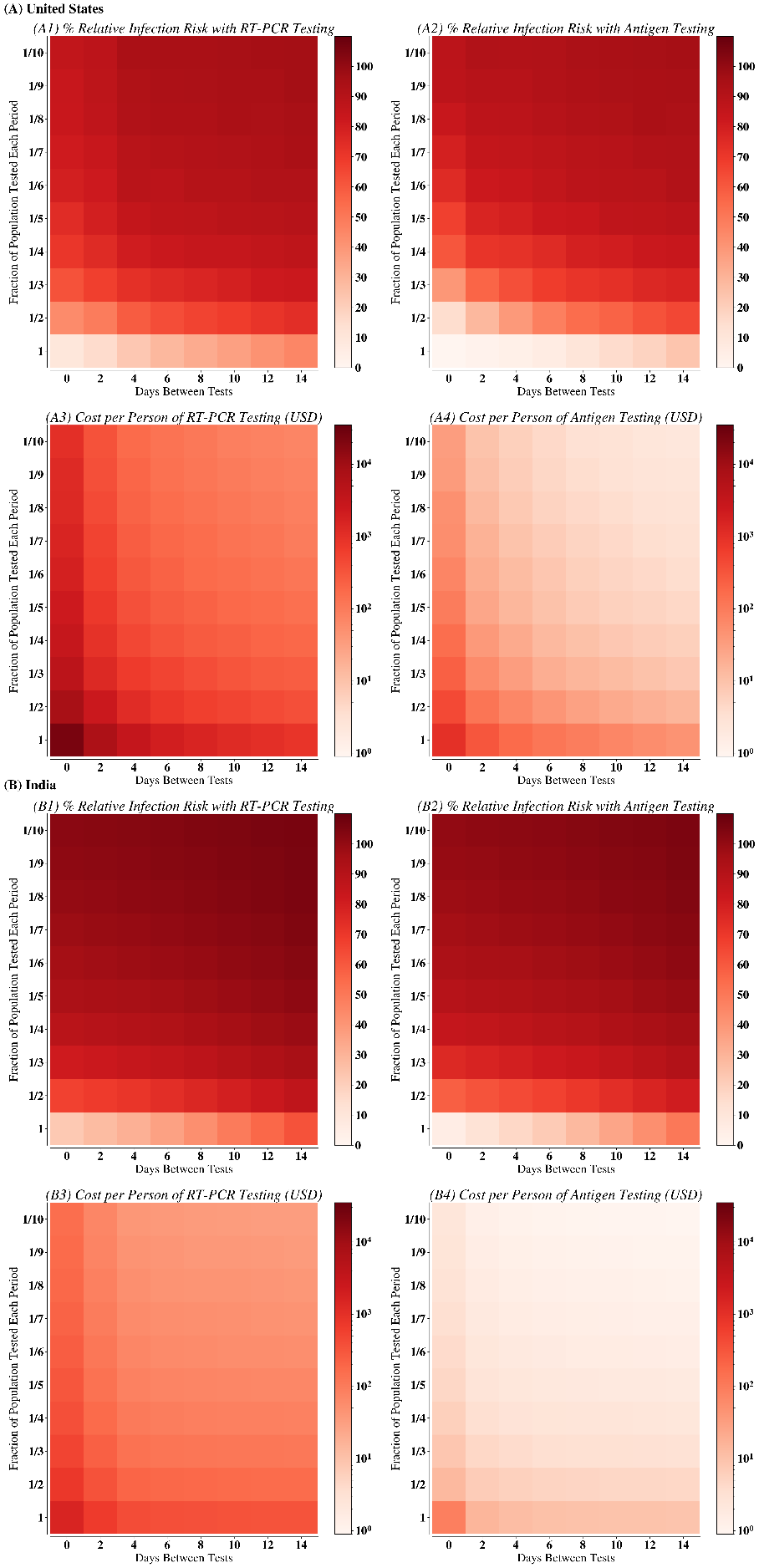

Figure S3: Complete values for limited data shown in Figs. 1 and 2.

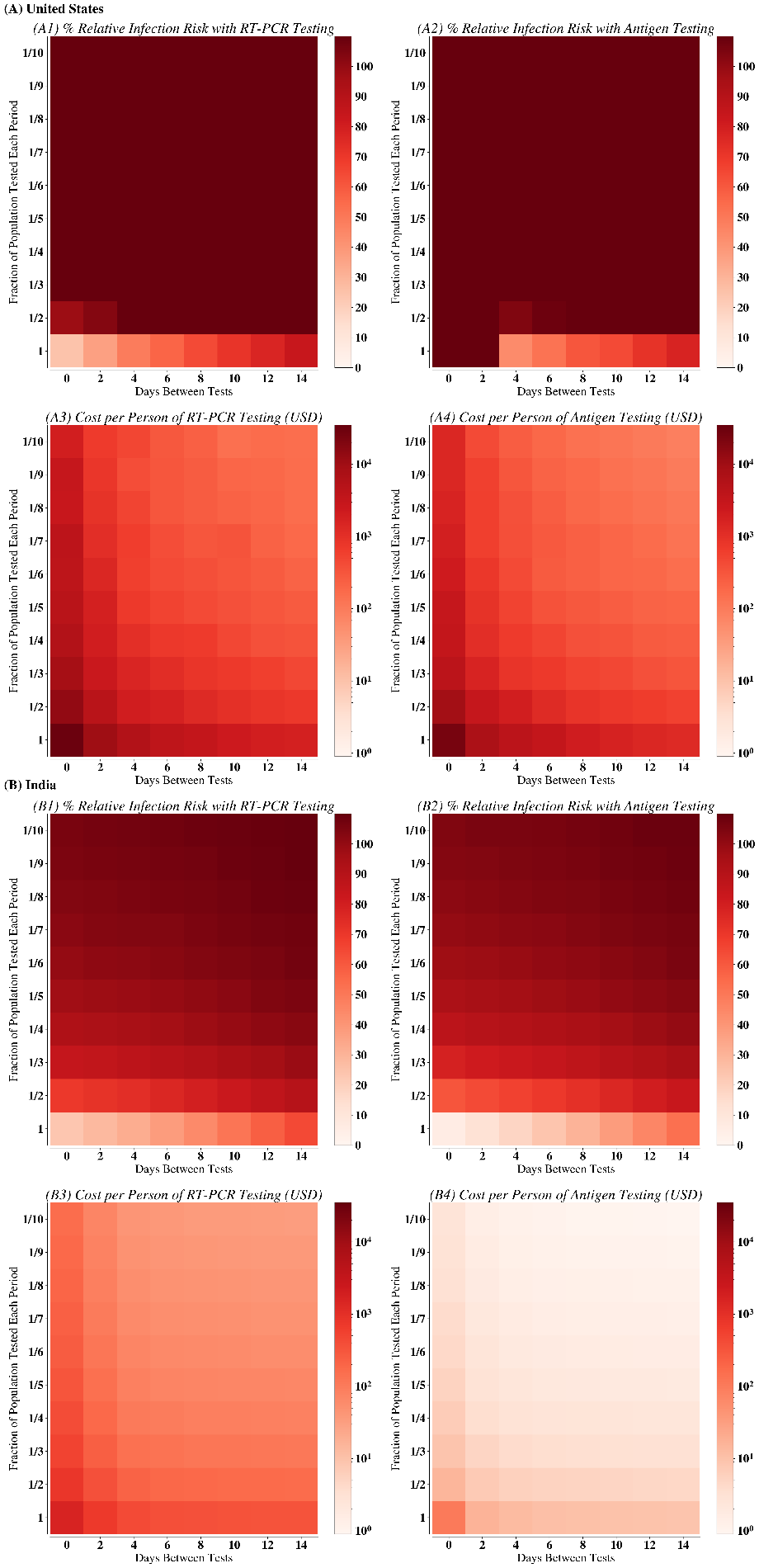

Figure S4: Upper bound of results shown in Fig. S3 and Figs. 1 and 1.

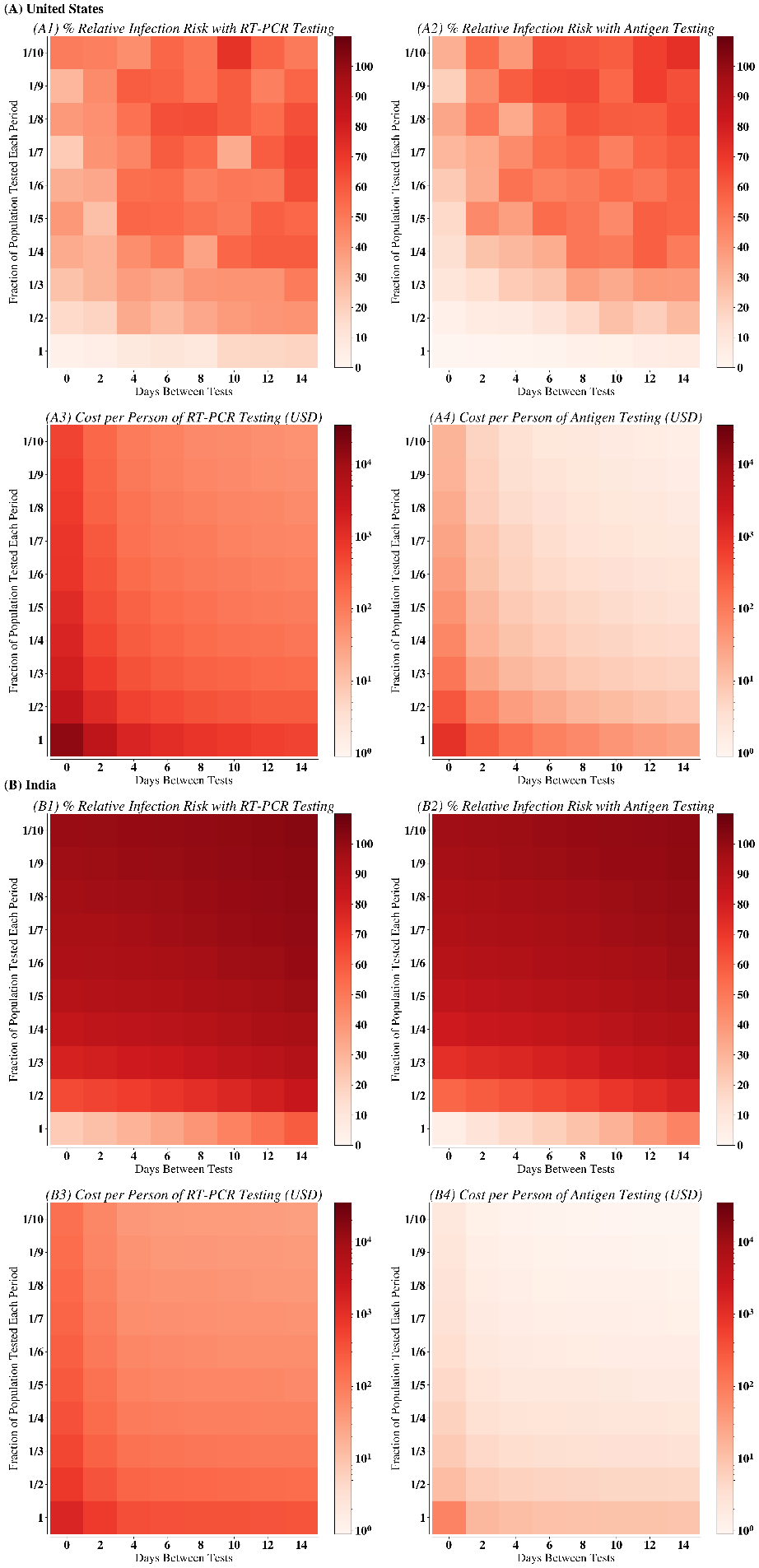

Figure S5: Lower bound on results shown in Fig. S3 and Figs. 1 and 1.

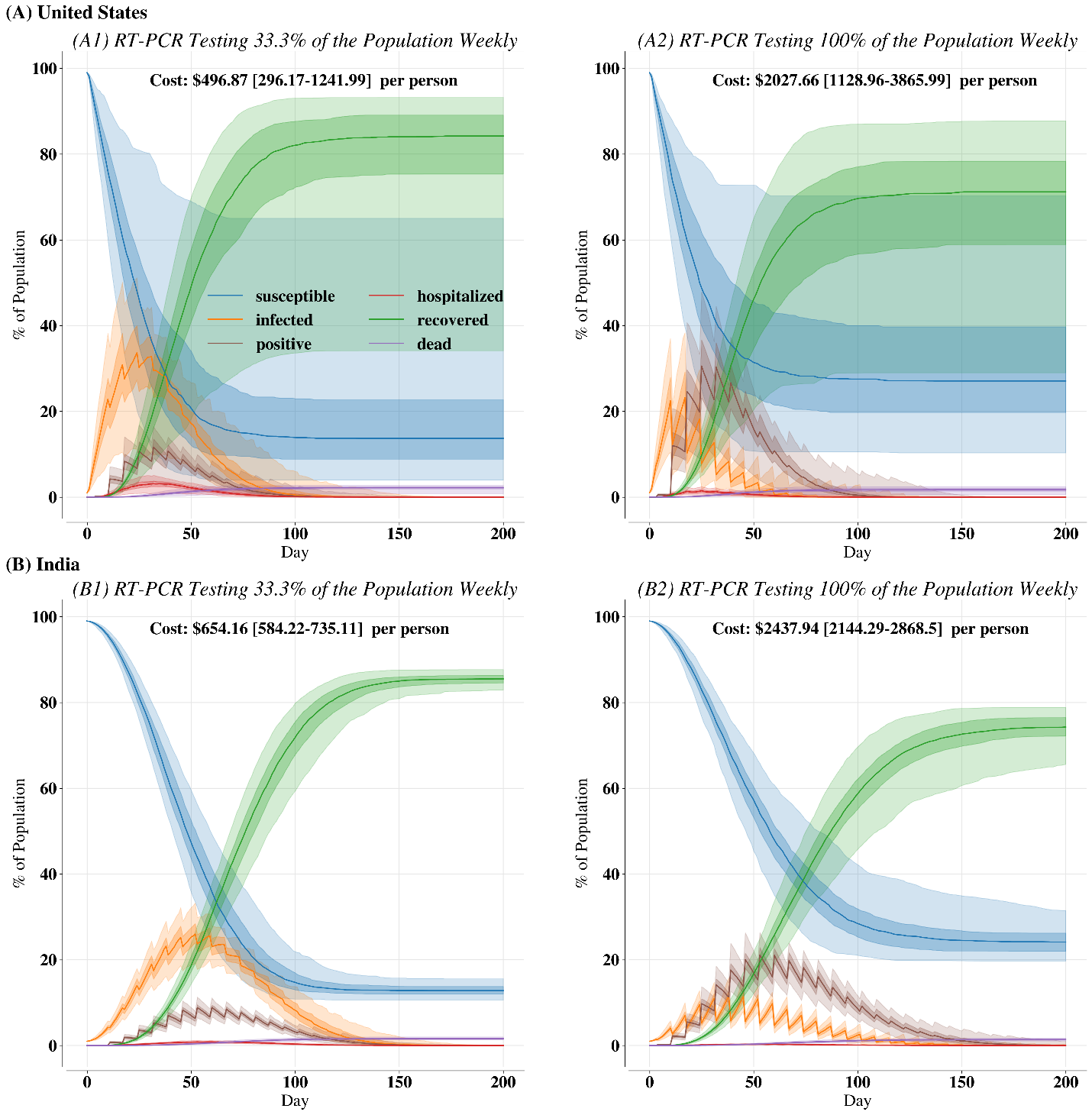

Figure S6: Exploratory RT-PCR testing scenarios for (A) United States and (B) India. (1) shows the disease course with 33.3% of the population tested one day once a week; (2) shows the disease course with 100% of the population tested one day once a week. Bold lines are the median over 200 independent replicates. Dark-shaded regions show 25^th^ to 75^th^ percentiles. Light-shaded regions are 2.5^th^ to 97.5^th^ percentiles. The cost estimates assume each individual without an infection is tested weekly. The sawtooth profile occurs because testing happens on a certain day and thus infected individuals are labelled as positive, resulting in a sudden decrease in infected individuals and an increase in positive individuals.

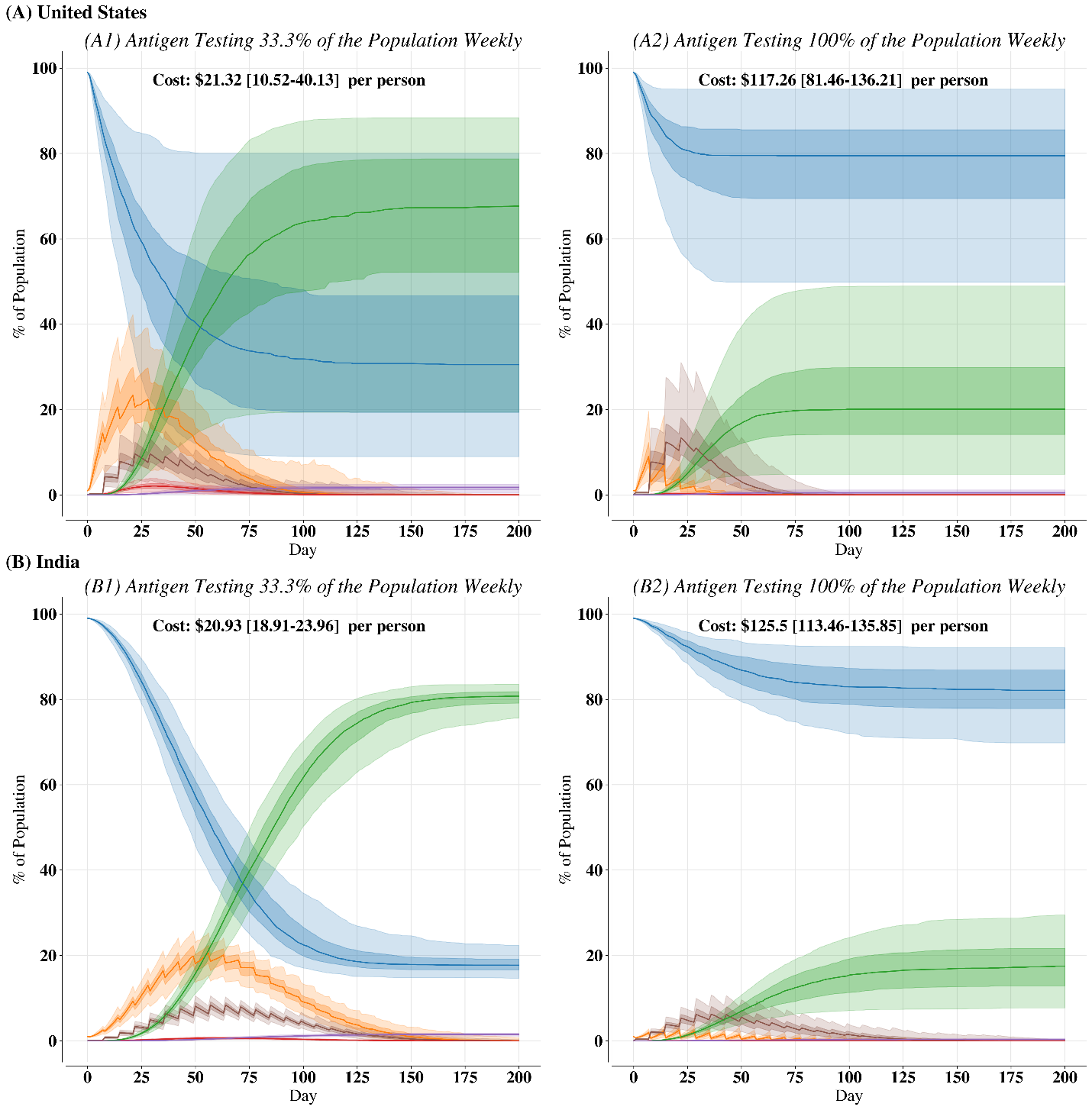

Figure S7: Exploratory antigen testing scenarios for (A) United States and (B) India. (1) shows the disease course with 33.3% of the population tested one day once a week; (2) shows the disease course with 100% of the population tested one day once a week. Bold lines are the median over 200 independent replicates. Dark-shaded regions show 25^th^ to 75^th^ percentiles. Light-shaded regions are 2.5^th^ to 97.5^th^ percentiles. The cost estimates assume each individual without an infection is tested weekly. The sawtooth profile occurs because testing happens on a certain day and thus infected individuals are labelled as positive, resulting in a sudden decrease in infected individuals and an increase in positive individuals.

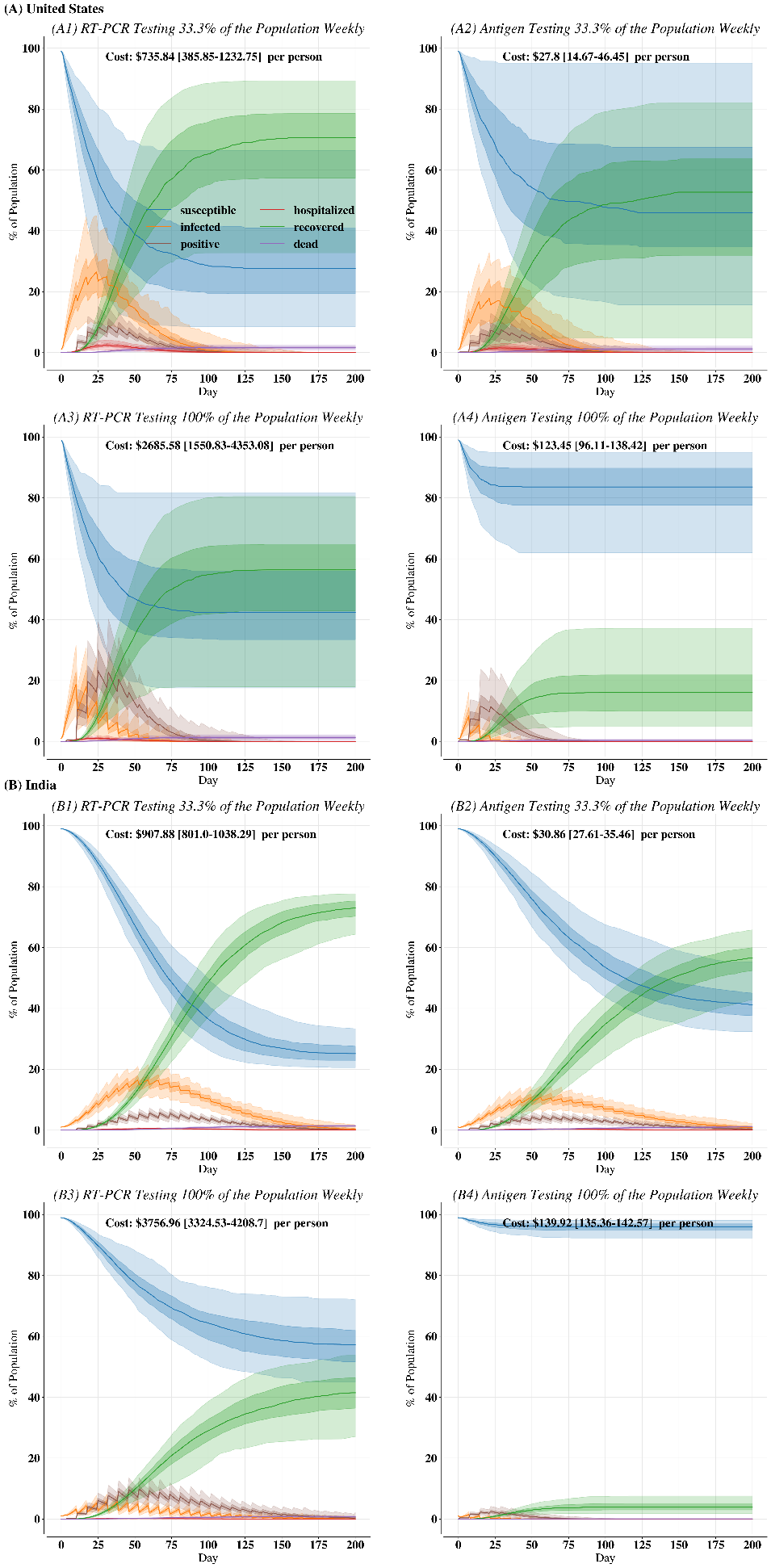

Figure S8: Exploratory testing scenarios in Fig. S7 with R_0_ = 2.

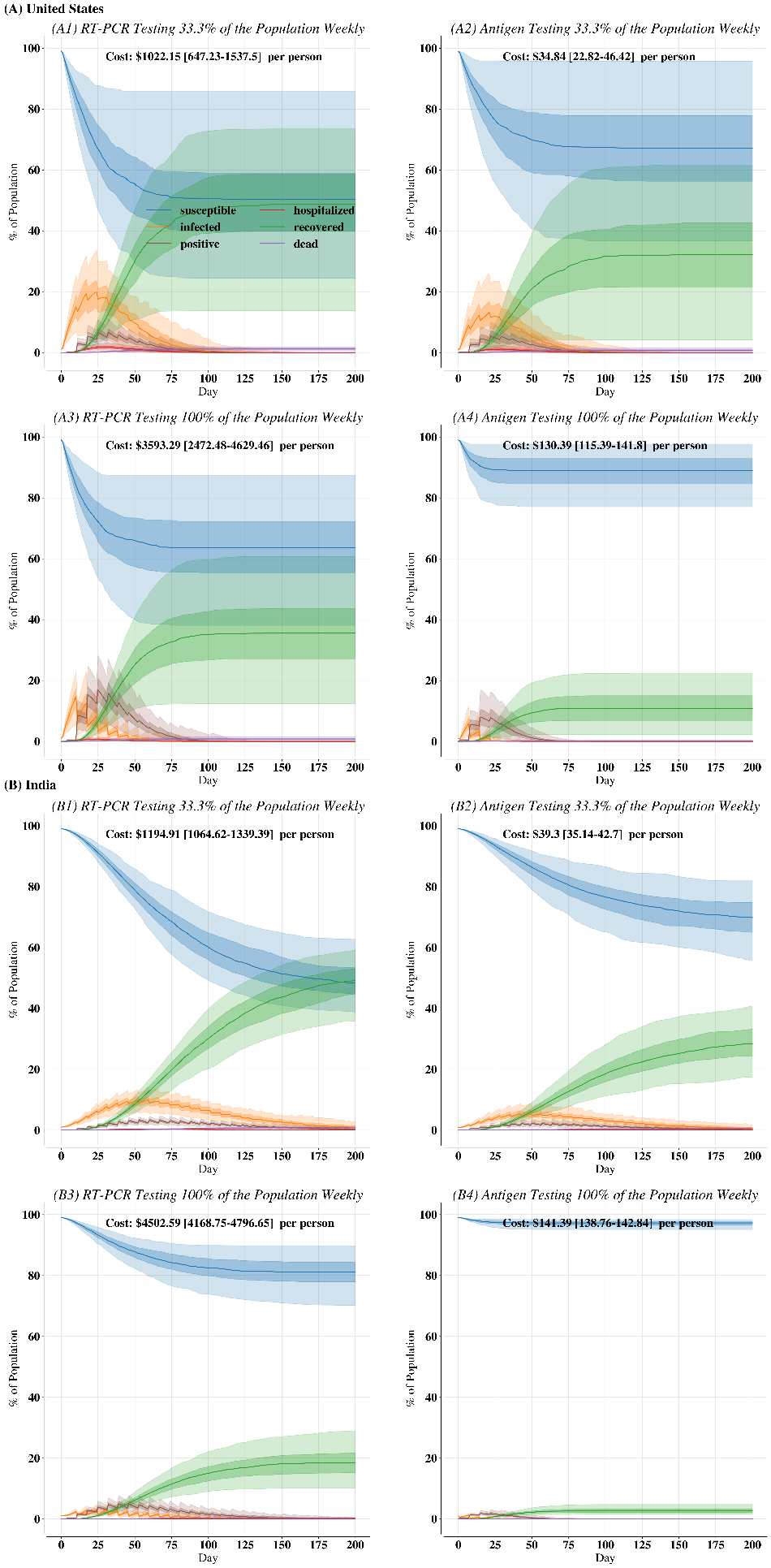

Figure S9: Exploratory testing scenarios in Fig. S7 with R_0_ = 1.5.

Table S2A: Standardized overall z-score regression results for predicting relative infection risk in each transmission setting by using testing at a certain coverage *g* (i.e., number of testing iterations required to surveil the full population), the frequency *f* (i.e., 1/(*d* + 1) where *d* is the number of days between testing occurrences), and the test used *r* (i.e., is the test used an RT-PCR assay?). The equation takes the form (A*g* + B*f*)/*gf +* C*r* where A, B, and C are the coefficients for *g, f,* and *r,* respectively*.* In this form, the numerator describes the risk of this testing procedure and the denominator standardizes the testing regiment by the characteristic time scale of the procedure; however, note that this equation can be arranged to be a linear function of 1/*f* and 1/*g*. Note that while Fig. 3 and 4 shows days between tests of 0, 4, 8, and 12 and fraction of population tested 1, ½, ¼, and 1/8, the regression equations are fit with 0, 2, 4, 6, 8, 10, 12, and 14 days between tests and 1, ½, 1/3, ¼, 1/5, 1/6, 1/7, 1/8, 1/9, and 1/10 as the fraction of the population tested.

|  | United States | India |
| --- | --- | --- |
| Coefficient for *g* | 0.2506 [0.2107 – 0.2906] | 0.1913 [0.1591 – 0.2235] |
| Coefficient for *f* | -0.9300 [-0.9700 – -0.8901] | -0.9578 [-0.9900 – -0.9256] |
| Coefficient for *r* | 0.1265 [0.0701 – 0.1830] | 0.0930 [0.0475 – 0.1385] |
| *R^2^* | 93.5% | 95.8% |
| F-statistic (p-val) | 763.1 (2.33e-93) | 1204. (4.44e-108) |
| AIC | 20.6872 | -48.42 |

Table S2B: Same results as in (A) but now for only RT-PCR testing. The equation takes the form (A*g* + B*f*)/*gf* where A and B are the coefficients for *g* and *f,* respectively.

|  | United States | India |
| --- | --- | --- |
| Coefficient for *g* | 0.2744 [0.2349 – 0.3139] | 0.1999 [0.1591 – 0.2408] |
| Coefficient for *f* | -0.9455 [-0.9850 – -0.9060] | -0.9629 [-1.0038– -0.9221] |
| *R^2^* | 96.9% | 96.6% |
| F-statistic (p-val) | 1232. (9.66e-60) | 1149. (1.36e-58) |
| AIC | -47.7118 | -42.2924 |

Table S2C: Same results as in (A) but now for only antigen testing. The equation takes the form (A*g* + B*f*)/*gf* where A and B are the coefficients for *g* and *f,* respectively.

|  | United States | India |
| --- | --- | --- |
| Coefficient for *g* | 0.2402 [0.1918 – 0.2886] | 0.1871 [0.1500 – 0.2243] |
| Coefficient for *f* | -0.9467 [-0.9951 – -0.8983] | -0.9684 [-1.0056– -0.9313] |
| *R^2^* | 95.3% | 97.2% |
| F-statistic (p-val) | 807.8 (7.37e-53) | 1397. (8.27e-62) |
| AIC | -15.2036 | -57.4782 |

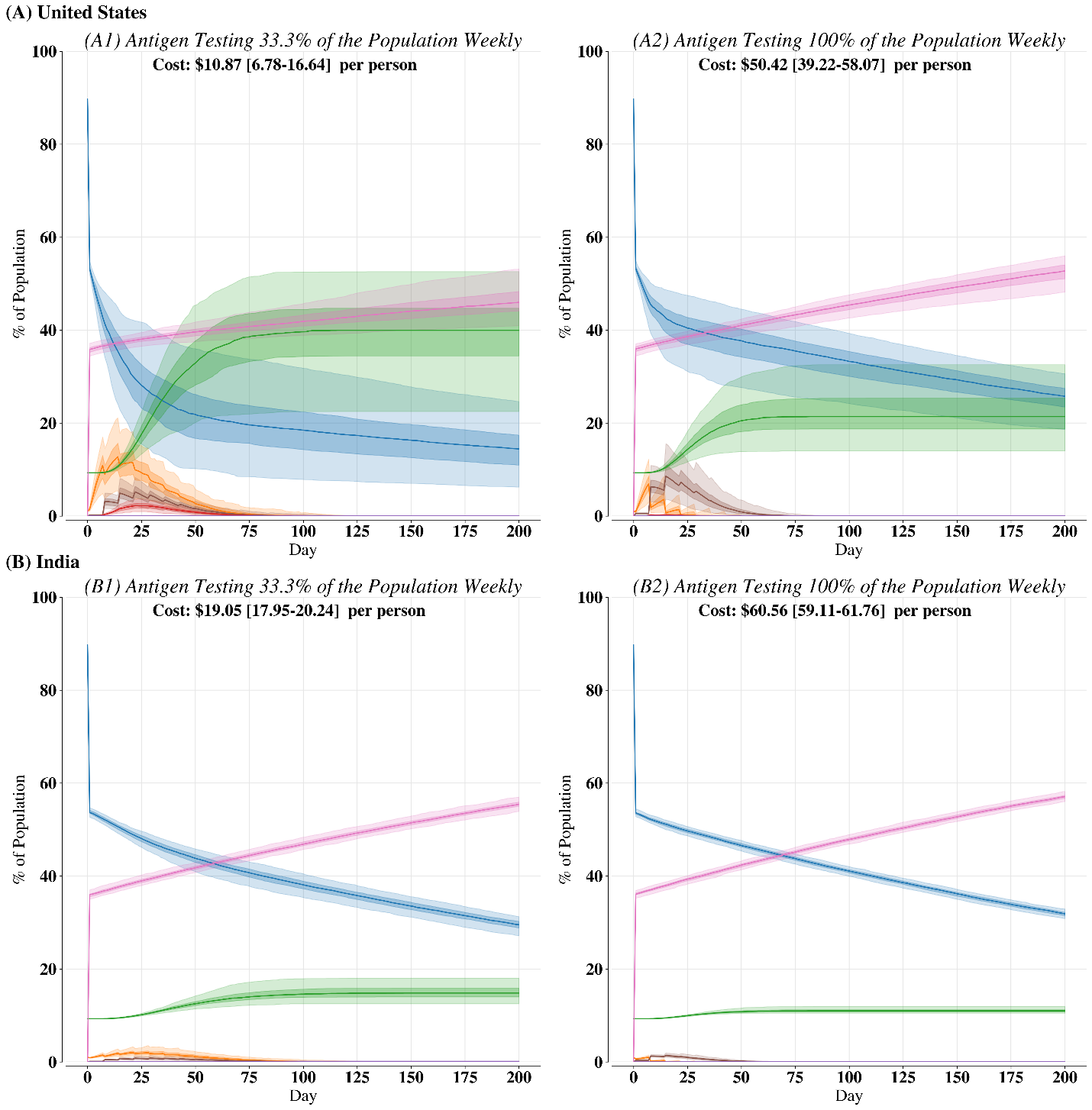

Figure S8: Disease course for (A) United States and (B) India with 40% of the population as initially vaccinated and then 0.25% of the susceptible population being vaccinated each subsequent day; vaccines are coupled with antigen testing (1) 33.3% or (2) 100% of the population weekly. Bold lines are the median over 200 independent replicates. Dark-shaded regions are the 25^th^ to 75^th^ percentiles. Light-shaded regions are 2.5^th^ to 97.5^th^ percentiles.

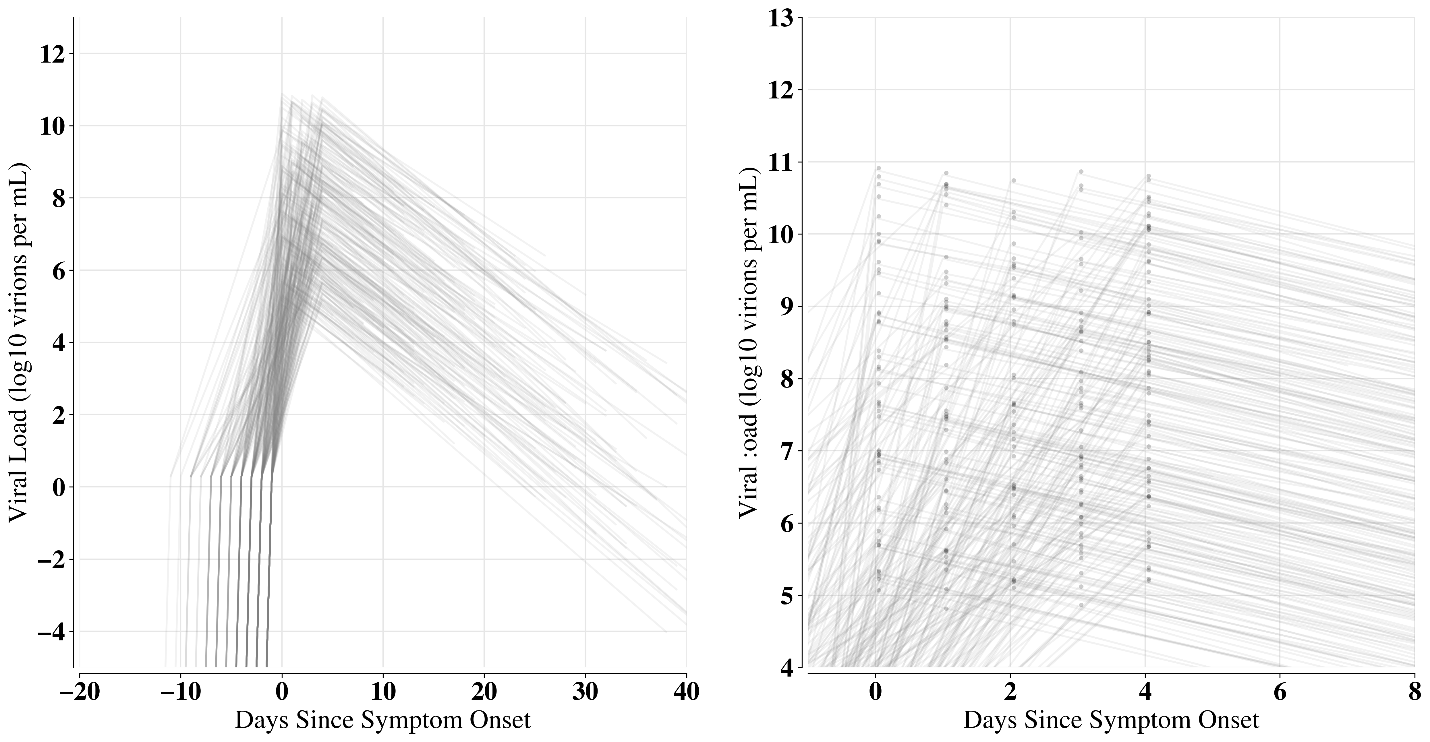

Figure S9: Viral load from a simple viral load kinetics model described in the Methods section. (Left) viral load over all days and (right) viral load zoomed in on days zero to four with dots on the peak viral load. Viral load increases linearly from negative infinity log10 virions per mL to a max from 5 to 11 log10 virions per mL that occurs on days zero through four. Log10 viral load then decreases linearly to the end of infection.

Table S3. Model Parameters

| Parameter | Value | Reference | Notes |
| --- | --- | --- | --- |
| R_0_ | 2.5 | US CDC National Center for Health Statistics | US CDC’s best guess |
| Incubation period | Lognormal with µ = 1.63, $\sigma= 0.5$ | McAloon et al. (2020) | From meta-analysis |
| Duration of infection given not hospitalized and recovered | Normal distribution with µ = 20.5, $\sigma$ = 6.7 | Hoertel et al. (2020) | Used in many modelling studies |
| Duration of infection prior to hospitalization: median (IQR) | 18-49 years: 6 (3, 10)  50-64 years: 6 (2, 10)  ≥65 years: 4 (1, 9) | US CDC National Center for Health Statistics | Estimates may change as trends change |
| Duration of infection given death: median (IQR) | 18-49 years: 15 (9, 25)  50-64 years: 17 (10, 26)  ≥65 years: 13 (8, 21) | US CDC National Center for Health Statistics | Estimates may change as trends change |
| Probability of admission to ICU given hospitalization: median (IQR) | 18-49 years: 23.8%  50-64 years: 36.1%  ≥65 years: 35.3% | US CDC National Center for Health Statistics | Estimates may change as trends change |
| Time spent in hospital: regular bed: median (IQR) | 18-49 years: 3 (2, 5)  50-64 years: 4 (2, 7)  ≥65 years: 6 (3, 10) | US CDC National Center for Health Statistics | Estimates may change as trends change |
| Time spent in hospital: ICU: median (IQR) | 18-49 years: 11 (6, 20)  50-64 years: 14 (8, 25)  ≥65 years: 12 (6, 20) | US CDC National Center for Health Statistics | Estimates may change as trends change |

Table S3 (cont.). Model Parameters

| Parameter | Value | Reference | Notes |
| --- | --- | --- | --- |
| Infectiousness over time | Shifted gamma distribution starting at -12.27, shape = 20.516, rate = 1.592 | He et al. (2020); corrected by Slifka et al. (2020) | The papers also provide uncertainties around these parameter estimates; we draw the parameters from the uncertainty distribution to construct the final gamma distribution. |
| Dispersion parameter for number of contacts infected | India: 0.51; US: 0.16 | Laxminarayan et al. (2020); Endo et al. (2020) |  |
| Testing sensitivities | RT-PCR: 100% viral load>0.91 log10 copies/mL  Antigen: 100% for viral load > 2 log10 copies/mL; 60% for viral load < 2 & >1 log10 copies/mL; 33.3% for viral load < 1 log10 copies/mL & > 0 log10 copies/mL; 26% for viral load < 0 log10 copies/mL  Antibody: 96% | Kleiboecker et al. (2020); Hirotsu et al. (2020); Stites and Wilen (2020) |  |
| Test specificities | RT-PCR: 95%  Antigen: 100%  Antibody: 95% | Stites and Wilen (2020); Hirotsu et al. (2020); Zhang et al. (2020) |  |
| Test costs | Antigen & antibody: $5, RT-PCR: $175 | Krouse and Abbot (2020) |  |

1.	Abbott, S. K. and B. What Kind of Covid Test Should I Get? Answers on Cost, Accuracy and More. *Wall Street Journal* (2020).

2.	Stites, E. C. & Wilen, C. B. The Interpretation of SARS-CoV-2 diagnostic tests. *Med* (2020).

10.	Zhang, Z., Hou, Y., Li, D. & Li, F. Diagnostic efficacy of anti‐SARS‐CoV‐2 IgG/IgM test for COVID‐19: A meta‐analysis. *J. Med. Virol.* (2020).
